# Cortical Lesion Expansion in Chronic Traumatic Brain Injury

**DOI:** 10.1101/2024.06.24.24309307

**Authors:** Holly J. Freeman, Alexander S. Atalay, Jian Li, Evie Sobczak, Natalie Gilmore, Samuel B. Snider, Brian C Healy, Holly Carrington, Enna Selmanovic, Ariel Pruyser, Lisa Bura, David Sheppard, David Hunt, Alan C. Seifert, Yelena G. Bodien, Jeanne M. Hoffman, Christine L. Mac Donald, Kristen Dams-O’Connor, Brian L. Edlow

## Abstract

Traumatic brain injury (TBI) is a risk factor for neurodegeneration and cognitive decline, yet the underlying pathophysiologic mechanisms are incompletely understood. This gap in knowledge is in part related to a lack of reliable and efficient methods for measuring cortical lesions in neuroimaging studies. The objective of this study was to develop a semi-automated lesion detection tool and apply it to an investigation of longitudinal changes in brain structure among individuals with chronic TBI. We identified 24 individuals with chronic moderate-to-severe TBI enrolled in the Late Effects of TBI (LETBI) study who had cortical lesions detected by T1-weighted MRI and underwent two MRI scans at least two years apart. Initial MRI scans were performed more than one year post-injury, and follow-up scans were performed 3.1 (IQR=1.7) years later. We leveraged FreeSurfer parcellations of T1-weighted MRI volumes and a recently developed super-resolution technique, SynthSR, to automate the identification of cortical lesions in this longitudinal dataset. Trained raters received the data in a randomized order and manually edited the automated lesion segmentations, yielding a final semi-automated lesion mask for each scan at each time point. Inter-rater variability was assessed in an independent cohort of 10 additional LETBI subjects with cortical lesions. The semi-automated lesion segmentations showed a high level of accuracy compared to “ground truth” lesion segmentations performed via manual segmentation by a separate blinded rater. In a longitudinal analysis of the semi-automated segmentations, lesion volume increased between the two time points with a median volume change of 4.91 (IQR=12.95) mL (p<0.0001). Lesion volume significantly expanded in 40 of 61 measured lesions (65.6%), as defined by a longitudinal volume increase that exceeded inter-rater variability. Longitudinal analyses showed similar changes in lesion volume using the ground-truth lesion segmentations. Inter-scan duration was not associated with the magnitude of lesion growth. Reliable and efficient semi-automated lesion segmentation is feasible in studies of chronic TBI, creating opportunities to elucidate mechanisms of post-traumatic neurodegeneration.

## Introduction

Traumatic brain injury (TBI) is a well-established risk factor for neurodegenerative diseases (Dams-O’Connor et al., 2020). The pathophysiologic mechanisms that link TBI to post-traumatic neurodegeneration (PTND) are not fully understood, though emerging evidence implicates a “polypathology” (Kenney et al., 2018) that includes axonal injury (Johnson et al., 2013), tau deposition (McKee et al., 2013), vascular injury (Dams-O’Connor et al., 2023; Sandsmark, Bashir, Wellington, & Diaz-Arrastia, 2019), and neuroinflammation (Johnson et al., 2013). An underexplored factor in the pathogenesis of PTND is the potential impact of focal cortical lesions, such as cerebral contusions, which are amongst the most common lesions in individuals with TBI (Vande Vyvere et al., 2024). It is unknown whether focal lesion size evolves during the chronic stage of TBI (i.e., more than one year post-injury) and whether this may contribute to clinical decline.

In addition to a paucity of longitudinal studies in individuals with chronic TBI, a key barrier to elucidating the impact of cortical lesions on PTND pathogenesis is methodological. Historically, lesions that disrupt the surface of the cerebral cortex have prevented MRI segmentation tools from parcellating the brain into its anatomic components (Merkley et al., 2008; Santhanam, Wilson, Oakes, & Weaver, 2019; Strangman et al., 2010). As a result, segmentation tools distributed with imaging analysis programs such as FreeSurfer (Fischl, 2012), FSL (Smith et al., 2004), and SPM (Friston et al., 1994) have been unable to robustly measure longitudinal lesion growth. Hence, patients with cortical lesions have typically been excluded from studies of cortical and subcortical volumetrics in individuals with TBI (Ding et al., 2008; Warner et al., 2010). Moreover, preliminary efforts at lesion segmentation have required substantial time by operators trained in human neuroanatomy (Diamond et al., 2020).

To address this methodological barrier and knowledge gap, we performed a longitudinal MRI study of individuals with chronic TBI and leveraged recent innovations in machine learning image analysis (Iglesias et al., 2023; Iglesias et al., 2021) to create a semi-automated lesion segmentation tool. We tested the ability of this semi-automated lesion segmentation tool to detect longitudinal changes in lesion volume in individuals with chronic TBI enrolled in the Late Effects of TBI (LETBI) study (Edlow et al., 2018). Our goal was to develop a tool that provides reliable and efficient measurement of cortical lesions to accelerate the study of PTND pathogenesis in individuals with chronic TBI.

## Methods

### Participant selection

Between 2014 and 2023, 305 participants were enrolled in the ongoing LETBI study at Mount Sinai School of Medicine (MSSM) and the University of Washington (UW) (Edlow et al., 2018). The LETBI study recruits individuals with a history of moderate or severe TBI. We use the United States Department of Defense classification of moderate TBI, which includes individuals considered by other classification systems as having “complicated mild” TBI (i.e., mild by Glasgow Coma Scale score criteria but with an intracranial lesion detected by brain imaging) (Defense, 2019). For the present longitudinal study, participants needed to have two MRI scans during consecutive study visits (≥2 years apart), each including T1-weighted (T1w) multi-echo magnetization prepared gradient-recalled echo (MEMPRAGE) scans (van der Kouwe, Benner, Salat, & Fischl, 2008) with a resolution of 1 mm isotropic.

Based on these criteria, 249 participants were excluded (n=220 not yet eligible for second study visit, n=29 without a T1w MEMPRAGE MRI dataset at both time points; see **Figure 1** for details). Of the n=220 excluded participants, scans from n=10 were randomly selected to form the inter-rater dataset. Of the remaining 56 participants, cortical lesions were identified in n=24 by a trained rater who visually inspected the T1w images. Lesions were defined by visible disruptions in the cortical grey matter or cortical grey/white junction. Lesions could extend into the adjacent subcortical white matter.

**Figure 1:**
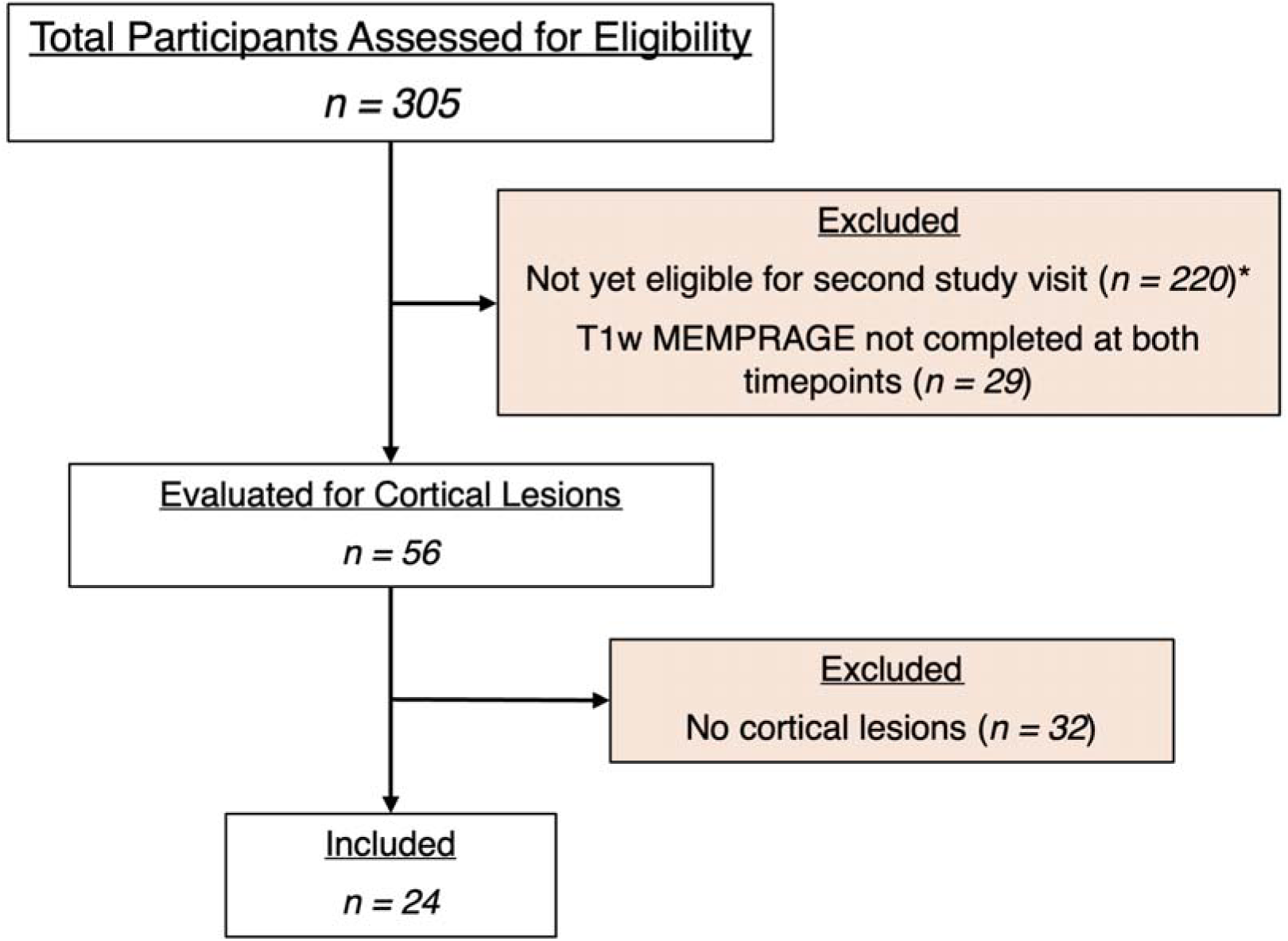
Evaluation of cohort for eligible participants. * For participants not yet eligible for a second study visit, 10 scans were assessed for cortical lesions to form an inter-rater dataset.

### Data acquisition, quality assessment, and processing

T1w images were obtained using Siemens Skyra, Philips Achieva, and Philips Ingenia Elition X scanners at 3 Tesla field strength. The images were acquired at 1 mm isotropic resolution. Siemens Skyra scans used a repetition time (TR) of 2,530 ms and echo times (TE) ranging from 1.79 ms to 7.37 ms. Philips Achieva scans used TRs ranging from 2,530 ms and TEs ranging from 1.67 to 7.07 ms. Philips Ingenia Elition X scans used a TR of 2,530 ms and a TE of 2.14 ms. Further information about the number of scans obtained from each scanner is provided in **Table 1**. Eleven participants underwent imaging on one scanner for their initial scan and a different scanner for their follow-up scan due to upgrades occurring during the study follow-up periods. Additional sequence parameters for the T1w sequences on each scanner have been previously reported (Edlow et al., 2018).

**Table 1.** MRI acquisition parameters for study participants.

| Manufacturer | Model | Field strength<br>(T) | TR/TE<br>(msec/msec) | Number of<br>scans |
| --- | --- | --- | --- | --- |
| Siemens | Skyra | 3T | 2530/1.79 – 7.37 | 18 |
| Philips | Achieva | 3T | 2530 / 1.67 – 7.07 | 11 |
| Philips | Ingenia Elition X | 3T | 2530 / 2.14 | 19 |

Qualitative and quantitative data quality assessments were performed on the processed images of all 24 subjects at both time points. Data uniformity and comparability across subjects and scanning platforms were examined, given the variations in sequence parameters. Visual quality assessments were based on the accuracy of FreeSurfer-generated surfaces (excluding those encompassing lesioned tissue) and the segmentation of subcortical structures, utilizing an accuracy rating scale adapted from Diamond et al. (2020). Signal-to-noise ratio (SNR) and contrast-to-noise ratio (CNR) were measured using the FreeSurfer tools ‘wm-anat-snr’ and ‘mri_cnr,’ calculating SNR in white-matter and the average of the WM-GM and GM-CSF contrasts, respectively. While no subjects were excluded due to quality assessment measures, differences were observed between the SNR distributions of enrollment sites, as reported in **Table 2**.

**Table 2.** Quantitative quality assessment of MRI data at each enrollment site.

| Enrollment site | Longitudinal cohort | SNR | CNR |
| --- | --- | --- | --- |
| MSSM | n=9 | 23.30 +/- 5.73 | 0.93 +/- 0.19 |
| UW | n=15 | 39.29 +/- 12.21 | 0.75 +/- 0.14 |

The T1w images were then processed, and the surfaces were constructed, using FreeSurfer v7.4 (Fischl, 2012). FreeSurfer processing involves motion correction, averaging of T1w images, removal of non-brain tissue, automated Talairach transformation, and segmentation of brain structures. It also includes intensity normalization, gray/white matter boundary tessellation, and topology correction. Further steps involve surface deformation, surface inflation, spherical atlas registration, cortical parcellation, and the creation of curvature and sulcal depth maps. To robustly segment neuroanatomic structures in brains with heterogeneous pathology, we used the Sequence Adaptive Multimodal SEGmentation (SAMSEG) tool (Cerri et al., 2021; Puonti, Iglesias, & Van Leemput, 2016), instead of the default automated segmentation (aseg) tool, before FreeSurfer recon-all. FreeSurfer reconstructions for all participants were completed successfully.

“Ground truth” lesion segmentation

Ground truth segmentations for all participants were established through manual tracing performed by a neurologist blinded to subject identification and time point. The process involved loading each T1w image into the FreeSurfer image viewer, Freeview. A blank label volume was created using the same geometry as the T1w image. The neurologist then manually segmented each lesioned area using the voxel edit tool, ensuring accurate and detailed delineation of the lesions. All segmentations were initially performed on a single label volume, which was later separated into unique values to indicate the presence of multiple lesions for individual subjects, thus creating a detailed ground truth segmentation volume at each time point for all participants.

### Semi-automated lesion segmentation

To minimize time requirements and reduce false negatives (i.e., missed labeling) in manual tracing, we developed a novel method for semi-automated lesion segmentation. As illustrated in **Figure 2A**, we leveraged SynthSR (Iglesias et al., 2023; Iglesias et al., 2021), a publicly available tool integrated within FreeSurfer that turns an MRI scan of any orientation, resolution, and contrast into a 1 mm isotropic T1w image while inpainting lesions.

**Figure 2:**
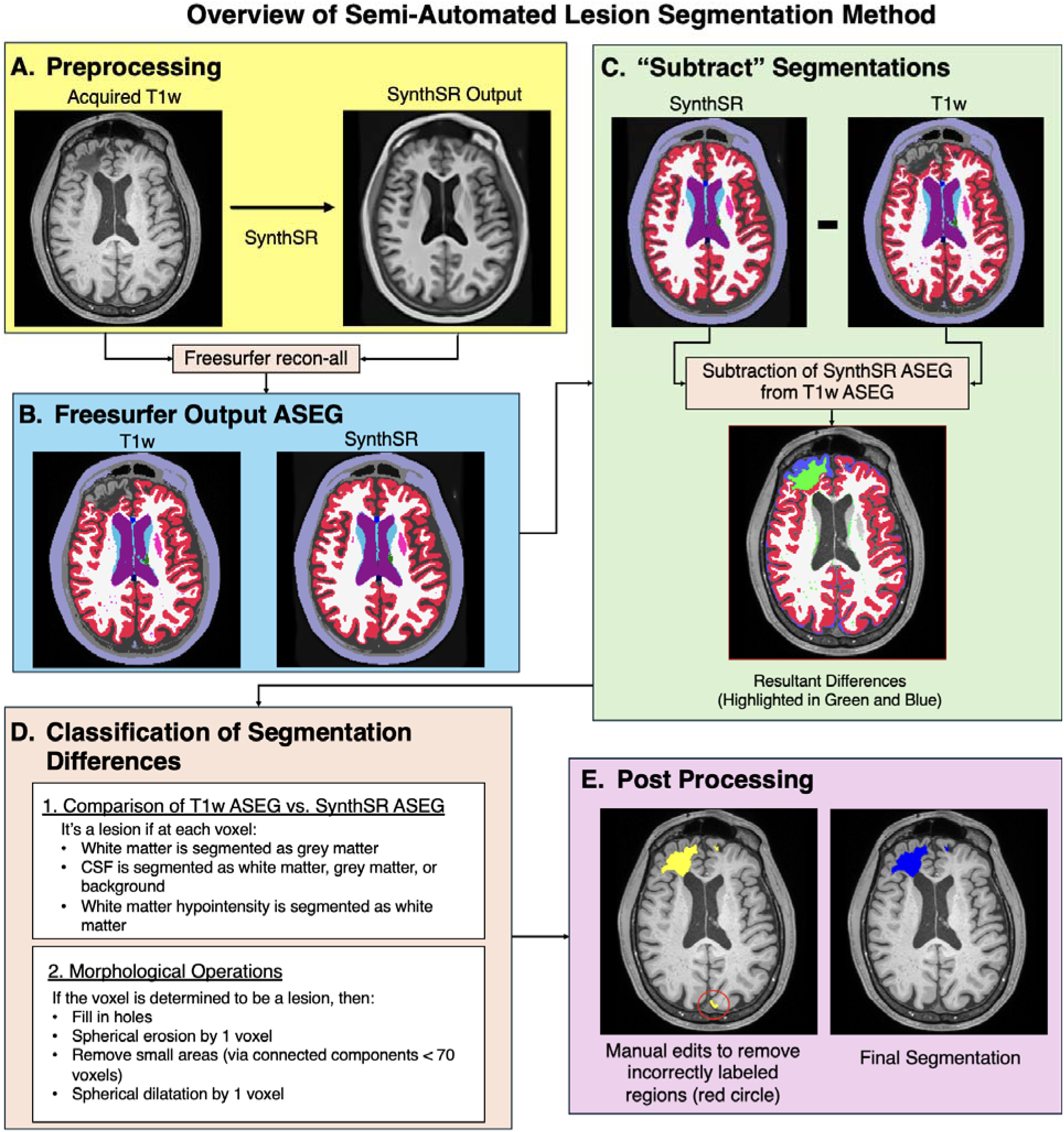
Overview of semi-automated lesion segmentation method. SynthSR images are generated for each acquired T1w image (A). Both images are then processed through Freesurfer recon-all, resulting in ASEG label volumes (B). The SynthSR ASEG is subtracted from the acquired T1w ASEG to highlight segmentation differences (C). These differences are classified voxel by voxel based on rules comparing the SynthSR and acquired T1w segmentation volumes. Identified lesions are then refined using morphological operations (D). Finally, the cleaned segmentation is reviewed for errors, including incorrectly labeled anatomy or missed lesions, and corrected to produce the final modified, semi-automated segmentation (E).

We applied SynthSR to T1w images for all participants and then repeated the FreeSurfer recon-all process on the synthesized images. We defined lesional voxels by comparing the SAMSEG (Cerri et al., 2023) labels from the synthesized image with those from the original T1w image (**Figure 2C**) using the following rules: a voxel is defined as a lesion if the segmentation label changed: 1) from white matter (in the original T1w recon) to gray matter (in the SynthSR recon); or 2) from CSF to background/white matter/gray matter; or 3) from white matter hypo-intensity to white matter. These rules were determined heuristically based on the segmentation label changes inside the lesional areas from a subset of our sample (n=5, randomly selected from the entire cohort and blinded to time point).

Subsequently, we applied morphological image processing (Soille, 2004) to remove false positives, reduce noise, and ensure that the detected lesional areas are topologically correct, including hole filling, spherical erosion/dilation, and area opening. Successful application of this pipeline facilitated the identification of clusters of lesioned voxels in the SynthSR-impainted volume, yielding an initial automated lesion segmentation mask (**Figure 2E**). A trained rater then performed manual edits (the only manual step in the semi-automated lesion segmentation method) to enhance the accuracy of lesion segmentation boundaries, yielding a final semi-automated lesion mask. In post-processing, this mask was separated into unique values to identify multiple lesions for a single subject. The rater performing manual edits for the semi-automated lesion segmentation method was blinded to the “ground truth” manual segmentations by the prior rater.

### Evaluation of inter-rater variability for the manual editing step of the semi-automated method

To determine inter-rater variability for the manual editing step of the semi-automated lesion segmentation method, we randomly selected 10 T1w images with lesions from the LETBI dataset that were not included in the 24-subject longitudinal dataset (i.e. subjects for whom there were not longitudinal data available). These 10 independent T1w images were edited by three raters, each of whom traced every lesion present on each scan. Raters were provided with SynthSR-generated segmentation masks (i.e., the initial automated lesion mask, as represented by the yellow lesion mask in **Figure 2E**) and instructed to revise the segmentations, creating a final semi-automated lesion mask (as represented by the blue lesion mask in **Figure 2E**). In post-processing, this mask was further separated into unique values to identify multiple lesions for individual subjects. The raters’ final lesion masks were then compared to measure inter-rater variability.

To test inter-rater variability, we performed Bland-Altman analyses for each pair of raters (Rater□1 vs. Rater□2, Rater□1 vs. Rater□3, and Rater□2 vs. Rater□3), calculating the mean difference (bias) and 95% limits of agreement (LOA). This analysis was completed on the volume of each lesion, with multiple lesions from patients contributing to the analysis and each lesion being treated as an independent observation. Additionally, we computed the Intraclass Correlation Coefficient (ICC) to assess the reliability of lesion-tracing. These analyses together assessed both agreement and reliability in lesion volumes, identifying any systematic biases or random variability. The LOA established a benchmark for subsequent statistical testing of longitudinal lesion expansion, allowing us to determine whether observed changes in lesion volume over time reflect lesion expansion or variability in the method.

### Comparison of semi-automated and ground truth segmentations

To evaluate the agreement of the methods, we compared edited semi-automated segmentations to the ground-truth segmentations (**Figure 3)** at both time points using Wilcoxon signed-rank tests and Bland-Altman analyses. The Wilcoxon tests assessed whether there were statistically significant differences in the volumes generated by the two methods, while the Bland-Altman analyses estimated the mean difference (bias), standard deviation, and limits of agreement between them.

**Figure 3:**
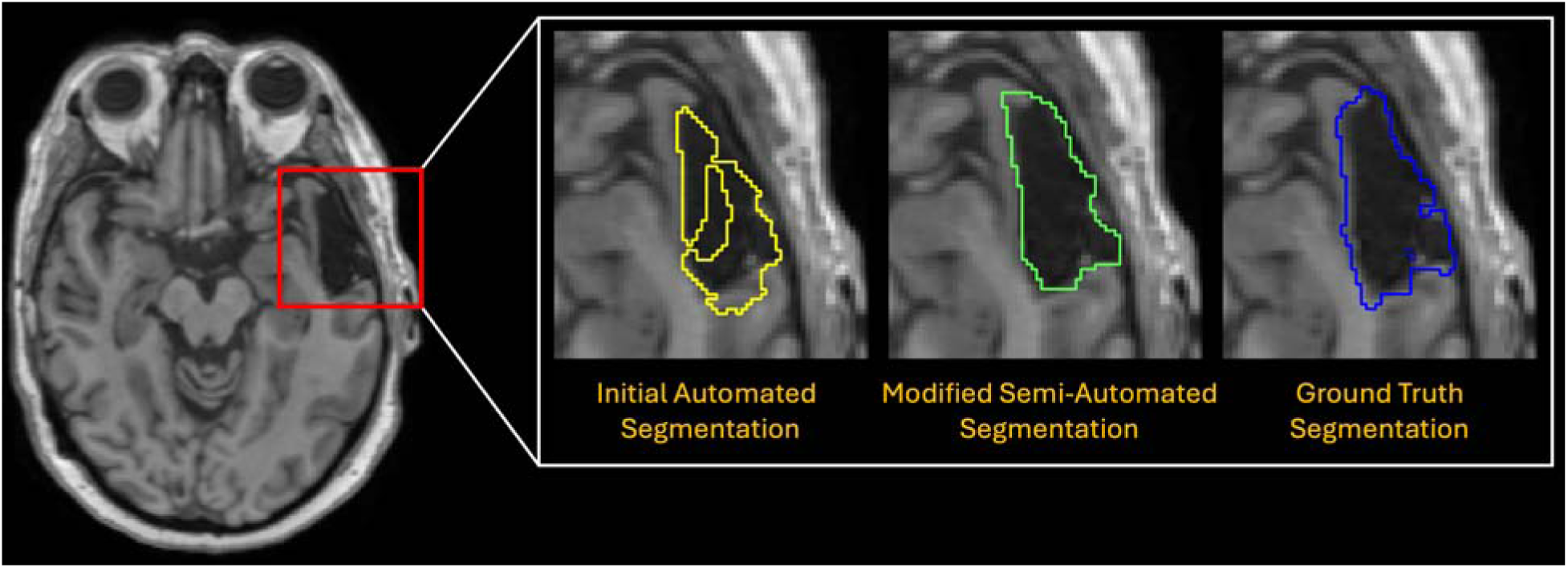
Comparison of the initial automated lesion segmentation (yellow), the modified semi-automated segmentation (green), which was revised by a trained rater, and the manually traced “ground truth” segmentation volume (blue).

### Testing for longitudinal changes in lesion volume

We hypothesized that there are detectable changes in lesion volume when comparing Visit 1 to Visit 2 for the entire cohort and when comparing single-subject changes in lesion volume to the null distribution of inter-rater variability (**Figure 4)**. We tested these two hypotheses using the semi-automated segmentations, as well as the ground truth segmentations. We began by comparing lesion voxel volumes, measured in mL, between Visit 1 and Visit 2, and then calculating the difference (Visit 2 - Visit 1) for each pair of measurements to determine the change in lesion volume. The statistical significance of these changes were assessed using the Wilcoxon Signed Rank test.

**Figure 4:**
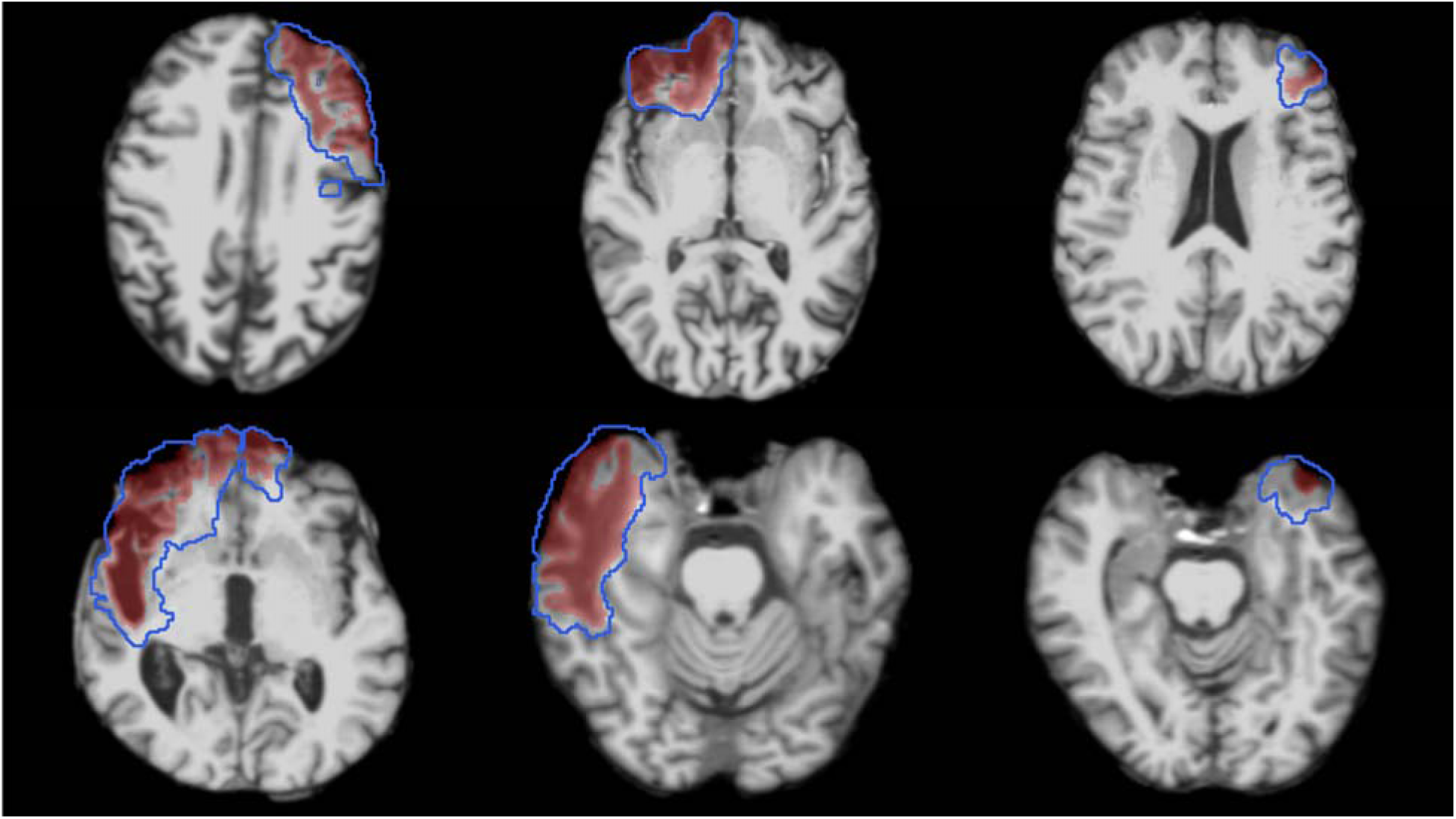
T1w images from Visit 1 of six representative subjects demonstrating the heterogeneous nature of lesion size and location. Images are overlayed with the ground truth lesion segmentations from visit 1 (red shaded regions) and visit 2 (blue outlined regions).

### Testing for changes in lesion volume compared to inter-rater variability

Next, we tested whether the observed longitudinal changes in Dice overlap and lesion volume exceeds the degree of inter-rater variability. Dice overlap and lesion volume differences were calculated for each lesion segmentation. For inter-rater data, Dice overlap and lesion volume differences were averaged across the three raters to generate composite scores for each lesion.

For the semi-automated and ground truth groups, dice overlap and lesion volume differences were derived by comparing Visit 2 to Visit 1 segmentations. Statistical significance of the differences between longitudinal changes in lesion measurements (i.e., Dice overlap and volume) and inter-rater variability was assessed using the Wilcoxon Rank Sum test, with a significance threshold of 0.05, to account for outliers and the small sample size. Finally, we operationally defined lesion expansion at the individual lesion level based on an increase in lesion volume greater than 1.5 times the SD over the mean of the inter-rater volume variability. We selected 1.5 SD as the cutoff based on the application of this statistical threshold to define abnormal cognitive performance in clinical practice (de Vent et al., 2020).

Longitudinal analyses of lesion volumes were performed in the subject’s native space at each time point. This method was selected instead of using the FreeSurfer longitudinal pipeline, which combines the two time points to generate a base image (Reuter, Schmansky, Rosas, & Fischl, 2012). The averaging process in the FreeSurfer pipeline would obscure the examination of lesion progression by blending the time points together, thus failing to capture dynamic changes in lesion size and location. By performing analyses in native space, we maintain the integrity of individual time point data, allowing for precise tracking of lesion growth and development over the study period without introducing registration artifacts.

### Evaluation of factors associated with lesion volume change

We examined the relationship between changes in lesion size (measured in mL) and the interval between imaging sessions (measured in days). We calculated the longitudinal change in lesion size for each subject by subtracting the lesion size at Visit 1 from that at Visit 2, separately for both the semi-automated and ground truth methods. Individual lesion clusters were matched between visits to directly compare changes over time. Pearson correlation coefficient (R) and two-tailed p-value were computed to assess the strength and significance of any linear relationship between changes in lesion size and duration between study visits. We applied Ordinary Least Squares (OLS) regression analysis to further investigate how age, sex and interval between study visits relate to changes in lesion volume.

## Results

### Patient and lesion characteristics

The 24 longitudinal participants ranged in age at Visit 1 from 33 to 73 years old, with a median age of 55.8 years (IQR = 14.3). Of these participants, nineteen were males. The 10 inter-rater participants ranged in age from 31 to 73 with a median age of 51.8 years (IQR = 22.8). Nine of the inter-rater participants were male. Additional descriptive statistics are provided in **Table 3**. Lesions were heterogeneous with respect to their neuroanatomic locations and were most prevalent in the anterior frontal and temporal lobes (**Figure 5**).

**Figure 5:**
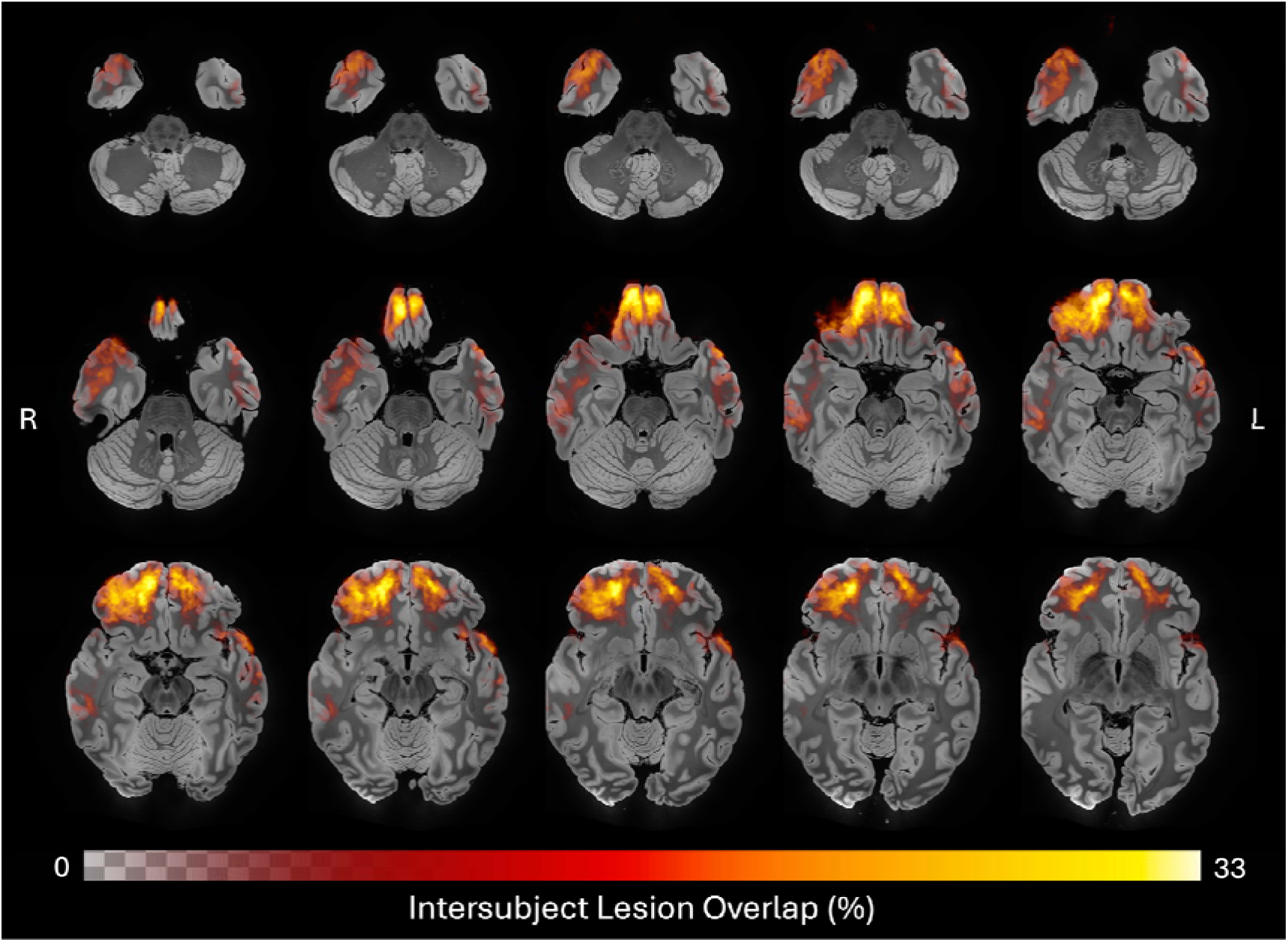
Neuroanatomic distribution of ground truth lesions across time points. Heatmap of all 48 ground truth lesion tracings registered to MNI space and overlayed on anatomical average, revealing a predominance of frontotemporal cortical lesions in this cohort. Color and opacity of the heatmap are modulated by the percent of lesion traces in each voxel, with the maximum overlap observed being 33%.

**Table 3.** Patient characteristics.

|  | <u>Longitudinal Dataset (n=24)</u> |  |  | <u>Inter-rater Dataset (n=10)</u> |  |  |
| --- | --- | --- | --- | --- | --- | --- |
|  | M | SD | Range | M | SD | Range |
| Age at First Visit (Years) | 55.8 | 11.2 | 33 - 73 | 51.8 | 14.5 | 31 - 73 |
| Visit Interval (Days) | 1328.5 | 498.7 | 734-2366 | - | - | - |
| Sex (Male:Female) | 19:5 |  |  | 9:1 |  |  |

### Inter-rater variability

We first assessed agreement among the three raters using Bland-Altman analysis (Table 4).

**Table 4.**
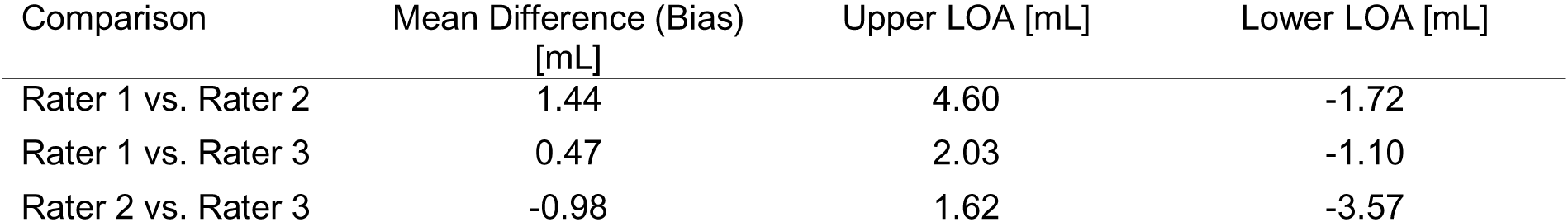
Inter-Rater Bland Altman Agreement Results.

| Comparison | Mean Difference (Bias)<br>[mL] | Upper LOA [mL] | Lower LOA [mL] |
| --- | --- | --- | --- |
| Rater 1 vs. Rater 2 | 1.44 | 4.60 | -1.72 |
| Rater 1 vs. Rater 3 | 0.47 | 2.03 | -1.10 |
| Rater 2 vs. Rater 3 | -0.98 | 1.62 | -3.57 |

The mean bias values between each pair of raters ranged from-0.98 mL to 1.44 mL, with LOA spanning from-3.57 mL to 4.60 mL. Additionally, the ICC was calculated to evaluate reliability among raters. The ICC (2,1) was 0.98 (95% CI: 0.93–0.99), indicating excellent reliability. These results demonstrate a high level of consistency across the raters, with minor differences likely attributable to individual rater preferences or the inherent complexity of lesion-tracing in chronic TBI. Variability within the LOA reflects the heterogenous and complex nature of the lesions, leading to differing identifications and tracings by raters.

### Semi-automated segmentation performance compared to ground truth segmentations

At Visit 1, the semi-automatic volume measurements yielded a mean bias of 2.42 mL (SD = 7.06 mL) relative to the ground truth measurements. The limits of agreement ranged from −11.41 mL to 16.25 mL, demonstrating a reasonably close alignment between the two methods. The Wilcoxon signed-rank test indicated no statistically significant difference (W = 276.00, p = 0.26), suggesting that the semi-automated method closely approximates the ground truth. At Visit 2, the semi-automated measurements showed a higher mean bias of −3.08 mL (SD = 9.71 mL) compared to the ground truth, with a wider range of agreement (−22.13 mL to 15.96 mL). This observation suggests that measurement differences varied more than in the first visit. A Wilcoxon signed-rank test revealed a significant difference between the two methods at this timepoint (W = 197.00, p = 0.019), suggesting that the discrepancies between the semi-automated and ground truth measurements were more pronounced at this visit.

Collectively, these findings at Visit 1 and Visit 2 suggest that the semi-automated method provides a reliable alternative to ground truth tracing, offering comparable accuracy and consistency, despite increased variability at the second timepoint (Figure 6). Although the time required for each segmentation method varied depending on lesion burden, we estimated that lesion adjustment using the semi-automated method required approximately 10–20 minutes per scan, compared to 60–90 minutes for manual segmentation.

**Figure 6:**
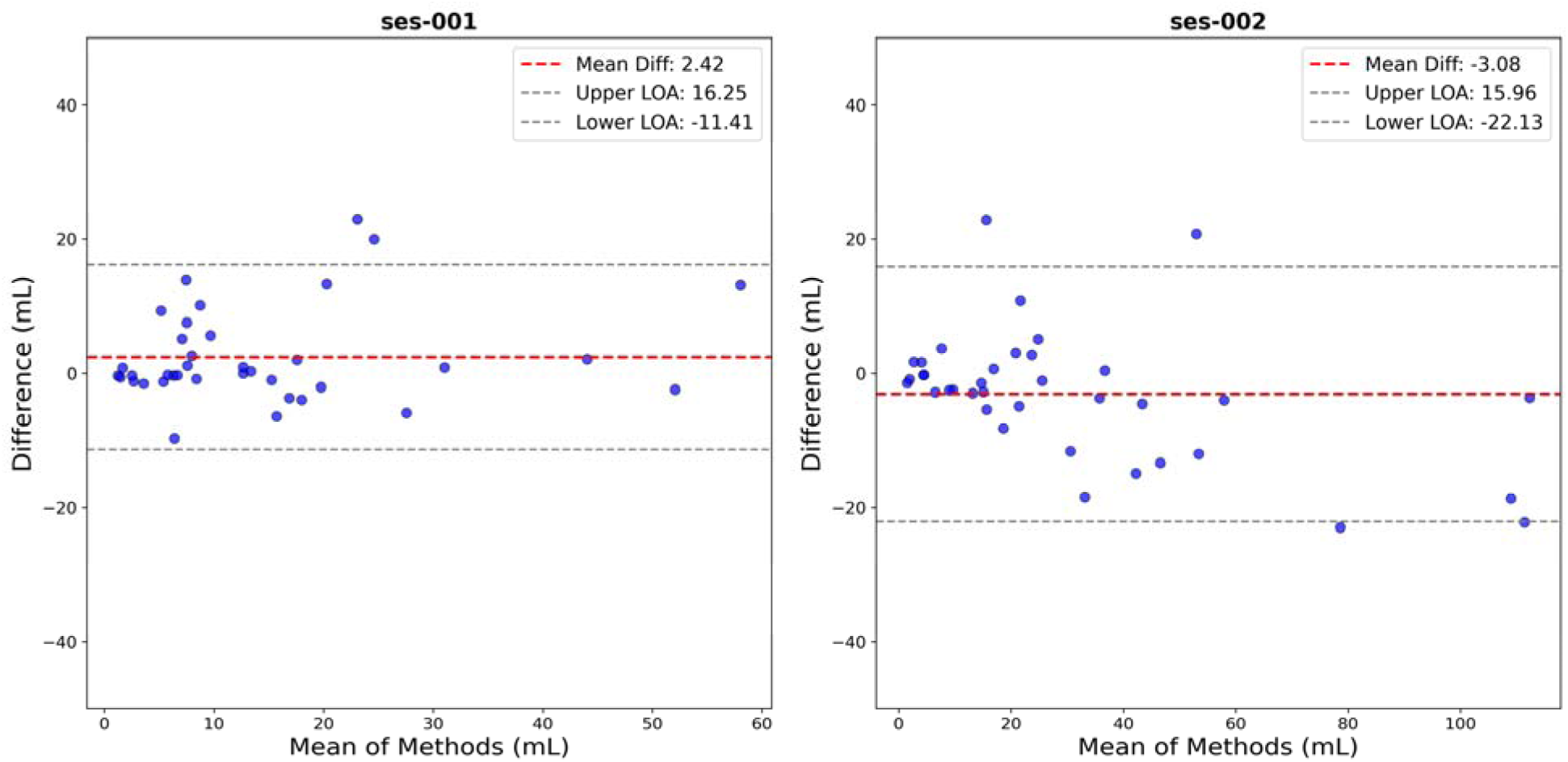
Bland-Altman agreement plots comparing lesion volume measurements across visits and methods. Each plot shows the difference versus the mean, with bias (dashed black lines) and limits of agreement (dashed red lines).

### Longitudinal changes in lesion volume

Longitudinal changes in lesion sizes derived from semi-automated segmentations at Visit 1 and Visit 2 ranged from-0.11 to 55.21 mL. The Wilcoxon signed-rank test results yielded a statistic of 1.0 at p < 0.0001, indicating an increase in lesion volume between Visit 1 and Visit 2.

Repeating the Wilcoxon signed-rank test using the ground truth segmentations similarly revealed an increase in lesion volume, ranging from 1.30 to 79.45 mL (W = 1.00, p < 0.0001).

### Changes in lesion volume compared to inter-rater variability

The longitudinal changes in Dice overlap from Visit 1 to Visit 2 exceeded inter-rater variability for both the semi-automated method (W = 1299.0, p < 0.0001) and the ground truth method (W = 684.5, p < 0.0001). Similarly, the increase in lesion volume from Visit 1 to Visit 2 exceeded inter-rater variability (i.e., the volume difference between raters for the same lesion) for both the semi-automated method (W = 225.0, p < 0.0001) and the ground truth method (W= 16.0, p < 0.0001) (**Figure 7)**. Further, 93.8% of lesions for the ground truth and 65.6% of lesions for the semi-automated experienced an increase in lesion volume greater than 1.5 times the SD over the mean of the inter-rater results (top dashed line in Figure 7, right).

**Figure 7:**
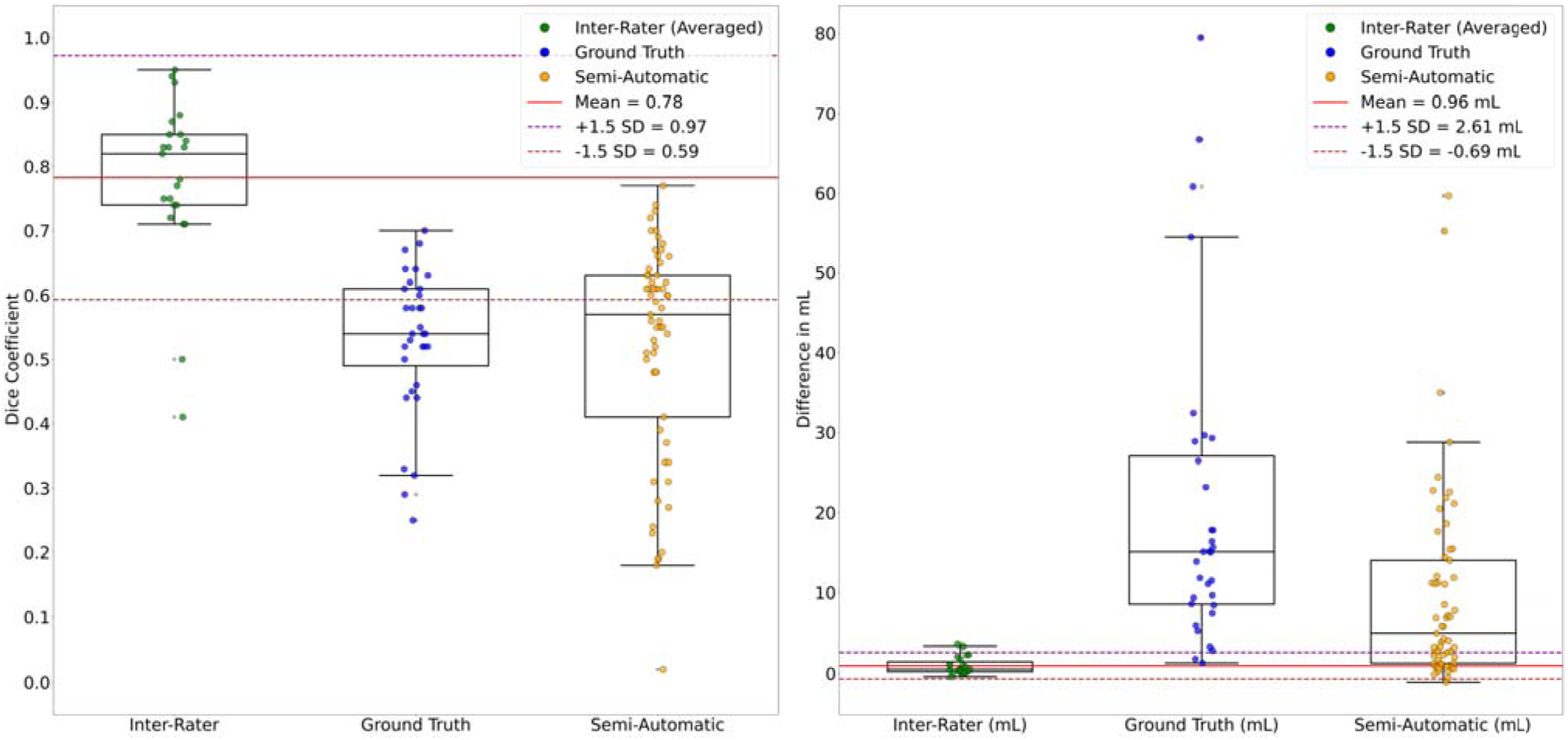
Comparison of Longitudinal Lesion Changes with Inter-rater Variability. The longitudinal changes in Dice scores (left panel) and volume measurements (right panel) from Visit 1 to Visit 2 are compared to their respective inter-rater reliability measures. The respective mean (red line) and 1.5 SD intervals (purple dashed lines) pertain to the inter-rater data.

### Lesion size variation across different visit intervals

Inter-visit intervals ranged from 727 to 2,366 days. Correlation analyses revealed no relationship between visit intervals and changes in lesion size for both the semi-automated (R =-0.12, p = 0.38) and ground truth (R =- 0.10, p = 0.58) methods (**Figure 8**). Regression models accounting for age, sex, and visit interval explained only 7.3% (R² = 0.073) and 5.2% (R² = 0.052) of the variance in lesion volume change, for the semi-automated and ground truth methods, respectively. After adjusting for the number of predictors, the adjusted R² values were 0.024 for semi-automated and-0.049 for ground truth, indicating minimal explanatory power. None of the individual predictors were statistically significant in either model (all p > 0.11)l. The overall F-statistics were 1.49 (p = 0.23) for semi-automated and 0.52 (p = 0.68) for ground truth, suggesting that the models did not effectively predict lesion volume changes. Together, these findings indicate that the observed changes in lesion size are not explained by age-related factors or influenced by sex, regardless of the measurement method used.

**Figure 8:**
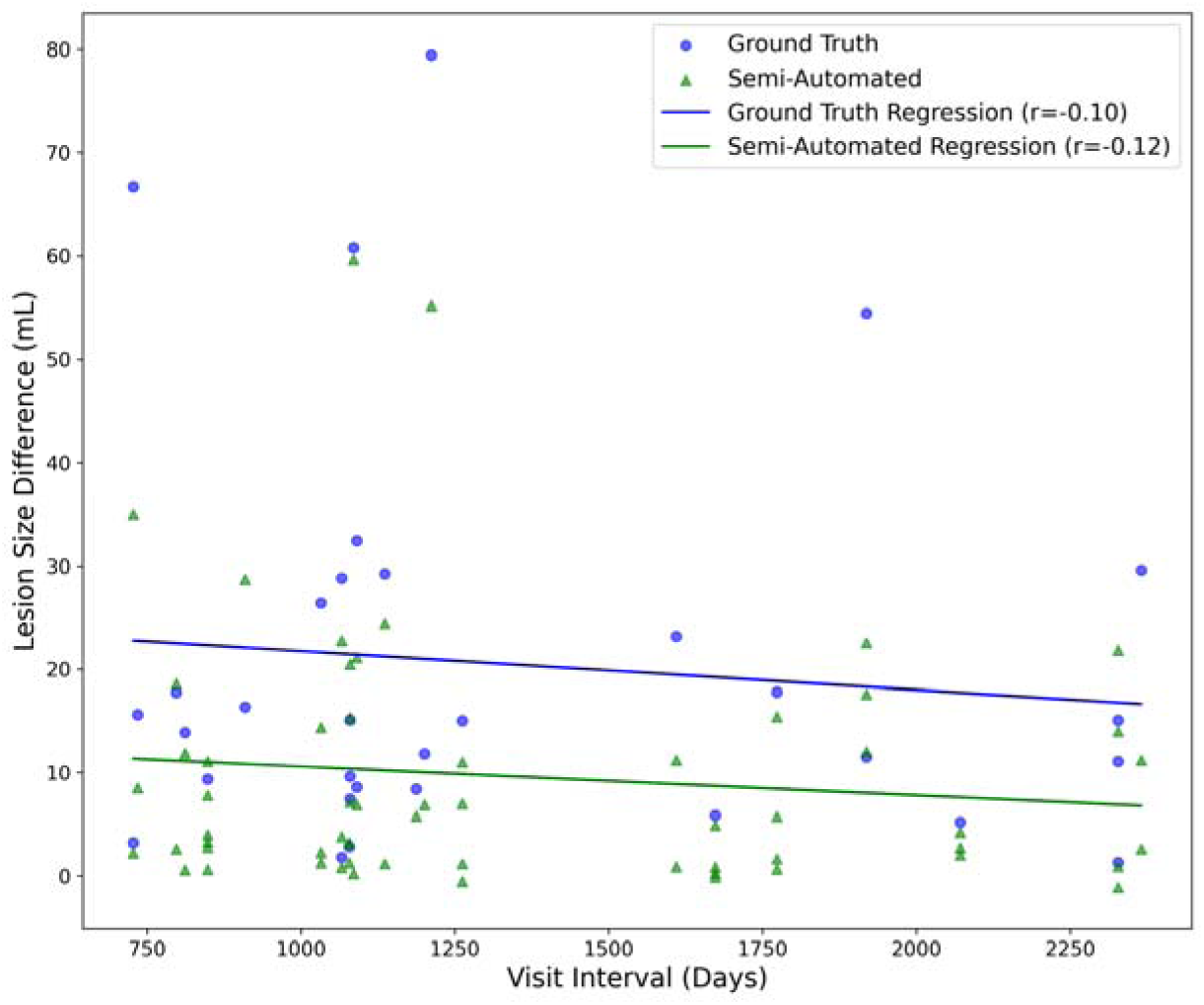
Correlation analysis examining the time between study visits and the calculated difference in lesion volume both for ground truth (blue) and semi-automated (green) methods.

## Discussion

In this longitudinal MRI study of 24 individuals with chronic TBI, we demonstrate the reliability and efficiency of a semi-automated cortical lesion segmentation tool. Our findings indicate that this FreeSurfer-based tool performs robustly against ground-truth manual tracings to segment lesions with improved time efficiency compared to previously developed methods (Diamond et al., 2020). Further, in a proof-of-principle application of the semi-automated lesion segmentation tool, we provide preliminary evidence that cortical lesions expand beyond one year post-injury, with 40 of 61 measured lesions (65.6%) experiencing lesion expansion on MRI scans performed at least 2 years apart. These observations raise the possibility that lesion expansion may contribute to PTND – a finding that will require confirmation in larger longitudinal studies with clinical-radiologic-pathological correlations. The semi-automated lesion tool thus creates new opportunities to investigate the role of cortical lesions in the pathogenesis of post-traumatic PTND.

The semi-automated lesion segmentation tool developed here builds upon recent innovations in machine learning-based imaging analysis, most notably SynthSR (Iglesias et al., 2023; Iglesias et al., 2021). What distinguishes this tool from previously developed lesion segmentation methods is: 1) increased efficiency when compared to traditional manual tracing; 2) improved accessibility and reproducibility; and 3) scalability for rapid, anatomically accurate lesion segmentation of large datasets. The new semi-automated tool demonstrates robust performance characteristics against “ground-truth” manual lesion segmentations, as evidenced by the strong agreement observed between the two methods at Visit 1. The observed discrepancies at Visit 2 may reflect a heightened sensitivity of the semi-automated method to more complex lesion morphologies, which could account for the increased variability in measurements. Collectively, these findings underscore the reliable performance of semi-automated segmentation compared to traditional manual tracing, but also the potential for future improvement in the reliability of the method.

Importantly, the semi-automated lesion segmentation tool requires human input to refine and optimize lesion boundaries – a step that reflects the inherent challenge of training automated tools to detect traumatic lesions, which often have heterogeneous signal characteristics related to variable distributions of gliosis, demyelination, and encephalomalacia. This manual editing step required approximately 10-20 minutes per MRI scan, and we estimate that several hours of training were required for each rater performing the manual editing. Nonetheless, the time required for the manual editing step in the newly proposed method is far less than for our previously published lesion segmentation method (Diamond et al., 2020). While the prior tool required manual creation of set points along the entire lesion surface, the new method requires only a small number of voxel-based edits in volumetric space. A key future direction will be to determine whether full automation is reliable. This goal may be attainable via integration with recently developed methods such as VoxelPrompt (Hoopes, Butoi, Guttag, & Dalca, 2024) and FastSurfer-LIT (Pollak, Kugler, Bauer, Ruber, & Reuter, 2025). We currently recommend manual editing of segmented lesions until further studies confirm the reliability of fully automated methods.

The lesion expansion observed in this cohort is consistent with and builds upon the growing evidence base indicating that pathological processes in TBI persist and progress in the chronic setting, even beyond one year post-injury. The clinical significance of lesion expansion is well established in the acute stages of care (Adatia, Newcombe, & Menon, 2021), wherein expansion of an acute lesion may cause mass effect and herniation. In the chronic stage, there are many unanswered questions about the underlying mechanisms and temporal dynamics of TBI lesion expansion. Prior evidence from histopathology (Johnson et al., 2013; Witcher et al., 2021) and neuroimaging studies suggests that inflammation persists in the chronic stage of TBI (Edlow et al., 2024; Scott et al., 2018), but whether chronic lesion expansion is attributable to inflammation, gliosis, microvascular ischemia, or some combination of factors is unknown. Elucidating mechanisms of chronic lesion expansion will require pathological-radiologic correlation analyses, which the LETBI study is designed to perform, given the premortem consent for autopsy provided by LETBI participants (Edlow et al., 2018). The absence of an association between lesion expansion and time between scans suggests that lesion expansion occurs at variable rates, though this preliminary observation will require future studies with larger sample sizes to confirm.

Several limitations should be considered when interpreting the results of this study. The small sample size of 24 individuals with chronic TBI limits the generalizability of our results, necessitating larger cohorts for validation. Only investigating two imaging time points and the relatively brief follow-up period of 3.5 +/- 1.2 years is also insufficient to elucidate the long-term trajectory of lesion expansion and its implications for PTND. The potential contribution of cortical lesion expansion to the pathogenesis of PTND is unknown and will require future studies with sufficiently large sample sizes to account for other risk factors, as well as protective factors. While the semi-automated tool improves efficiency, it still requires manual input for refining lesion boundaries, introducing potential variability and subjectivity. Additionally, the heterogeneous neuroanatomic locations and signal characteristics of traumatic lesions further complicate the segmentation process, as the tool may not uniformly handle all types of lesions with the same accuracy. Lastly, this study did not test for cognitive and functional correlates of lesion expansion – a crucial area for future research. Addressing these limitations will be essential for advancing our understanding of cortical lesion dynamics in chronic TBI.

In summary, we developed and implemented a semi-automated lesion detection tool that accurately identifies and efficiently quantifies the volume of cortical lesions in individuals with chronic TBI. Further, we provide proof-of-principle evidence that this lesion segmentation tool can detect longitudinal lesion growth in the chronic stage of TBI. Future applications of this tool have the potential to elucidate the pathophysiologic links between lesion expansion and the clinical expression of PTND, including in individuals with TBI resulting in large cortical lesions that would otherwise exclude them from analyses of neuroimaging data. Ultimately, the integration of lesion segmentation into clinical MRI workflows has the potential to inform preventive, diagnostic, prognostic, and therapeutic strategies for individuals with chronic TBI.

## Author Contributions

HJF, KD-O’C and BLE conceived and designed the study, and wrote the original draft of the manuscript. HC, ES, AP, LB, DS, DH, ACS, JMH, and CLM acquired the data.

HJF, ASA, JL, ES, NG, SBS, YGB, CLM, and BLE processed and analyzed the imaging data. HJF, ASA, ES, NG, SBS, and BLE performed the lesion tracings. HJF, JL and BCH performed the statistical analyses. CMD, KD-O’C and BLE contributed to project administration and supervision. All authors contributed to the editing of the manuscript.

## Data Availability

The imaging data are available upon reasonable request.

## Data Availability

Data are available upon reasonable request to the authors.

